# Defining predatory journals and responding to the threat they pose: a modified Delphi consensus process

**DOI:** 10.1101/19010850

**Authors:** Samantha Cukier, Manoj M. Lalu, Gregory L. Bryson, Kelly D. Cobey, Agnes Grudniewicz, David Moher

**Author notes:** **Corresponding author** David Moher, Centre for Journalology, Clinical Epidemiology Program, Ottawa Hospital Research Institute, Ottawa, ON, Canada K1H 8L6, (x79424). **Author contributions (CReDiT):** Conceptualization: KC, ML, DM Methodology: KC, ML, AG, DM Project administration: ML, GB, DM Investigation (data collection): SC Writing – Original Draft: SC Writing – Review & Editing: All authors Supervision: DM.

## Abstract

**Background:** Posing as legitimate open access outlets, predatory journals and publishers threaten the integrity of academic publishing by not following publication best practices. Currently, there is no agreed upon definition of predatory journals, making it difficult for funders and academic institutions to generate practical guidance or policy to ensure their members do not publish in these channels.

**Methods:** We conducted a modified three-round Delphi survey of an international group of academics, funders, policy makers, journal editors, publishers and others, to generate a consensus definition of predatory journals and suggested ways the research community should respond to the problem.

**Results:** A total of 45 participants completed the survey on predatory journals and publishers. We reached consensus on 18 items out of a total of 33, to be included in a consensus definition of predatory journals and publishers. We came to consensus on educational outreach and policy initiatives on which to focus, including the development of a single checklist to detect predatory journals and publishers, and public funding to support research in this general area. We identified technological solutions to address the problem: a ‘one-stop-shop’ website to consolidate information on the topic and a ‘predatory journal research observatory’ to identify ongoing research and analysis about predatory journals/publishers.

**Conclusions:** In bringing together an international group of diverse stakeholders, we were able to use a modified Delphi process to inform the development of a definition of predatory journals and publishers. This definition will help institutions, funders and other stakeholders generate practical guidance on avoiding predatory journals and publishers.

## INTRODUCTION

Predatory journals pose a serious threat to legitimate open access (OA) journals and to the broader scientific community^1^. They pose as authentic OA journals, however, they often fail to follow usual publication best practices, including peer review and editorial oversight^2^. These journals have self-interest as a goal, and are often motivated to accept as many articles as possible to profit from article processing charges (APCs) which are common at OA journals. It is becoming increasingly difficult to distinguish articles published in predatory journals from legitimate journals as predatory journals are also finding their way into trusted sources like PubMed^3^.

Despite increasing attention to the problem of predatory publishing^4–8^, there is no agreed upon definition of what constitutes a predatory journal^9^. The absence of a consensus and operationalized definition makes it difficult to accurately identify and evaluate the problem. Without a definition, funders and academic institutions struggle to generate practical guidance or policy to ensure their members do not publish in predatory journals. Without appropriate attention to the problem of predatory publishing, the quality of scholarly communication is at risk; this includes the risk to researchers, academic institutions, and funders whose credibility may be questioned, and/or patients who will have given of their time in hopes of improving interventions or treatments, when in all likelihood this data would not be used^4^.

This paper is part of a program of scholarship that aims to establish a consensus definition of predatory journals and publishers, and establish ways in which the research community should respond to the problem. Cobey and colleagues^9^ reported on the first stage of the program, which was a scoping review to identify possible characteristics of predatory journals. Authors found that no consensus definition existed and there was a great deal of heterogeneity in the characteristics found. In this, the second stage of the research program, we used the characteristics identified from the scoping review to generate a consensus definition of predatory journals and also suggested ways the research community should respond to the problem.

Here we present details of this modified three round Delphi consensus study. A related paper, describing the consensus statement reached on predatory journals is described elsewhere^10^.

## METHODS

Prior to commencing this study, a protocol was drafted (https://osf.io/z6v7f/) and approved by the Ottawa Hospital Research Ethics Board (Ottawa, Ontario, Canada, 20180927-01H) (https://osf.io/ysw3g/). The protocol was posted on the Open Science Framework prior to initiating the study.

The Delphi method is a structured method to elicit opinions on given questions from a group of experts and stakeholders^11,12^. It is especially useful when exact knowledge is not available. The participants respond anonymously to questionnaires that sequentially incorporate feedback and are refined. Following each round, average group responses are provided to each respondent, allowing them to reconsider their own views on the topic. This is generally performed through electronic survey, however, for our modified Delphi the final round was held through a face-to-face meeting.

### Delphi Survey Questions – Predatory Journals and Publishers

The Delphi survey was made up of 18 questions and 28 sub-questions (see Appendix A). Questions encompassed three themes: (1) predatory journal definition; (2) educational outreach and policy initiatives on predatory publishing; and (3) developing technological solutions to stop submissions to predatory journals and other low-quality journals.

Questions for the first theme were informed by work identifying salient features of predatory journals^2^ as well as a scoping review of characteristics of probable predatory journals^9^. Questions for the remaining two themes were developed iteratively by members of the research team. The survey was reviewed by one individual external to the research team and then pilot tested by four others, including the one individual who reviewed the survey. Feedback received during review and piloting was incorporated into the survey.

### Modified Delphi Process

We used online Survey Monkey software (http://surveymonkey.com) to deliver rounds 1 and 2 of our Delphi survey electronically. Participants were invited via an email which included key information about the study, its purpose and how it would inform a consensus definition of predatory journals and directions for future research. Rounds 1 and 2 were available online for three weeks each. Two reminders were sent to participants at day seven and fourteen. Round 3 was conducted at our Predatory Summit, using Poll Everywhere software (http://www.polleverywhere.com), where participants could respond to survey questions through live polling, watch results, and participate in a face-to-face discussion.

For each of the questions, participants were asked to respond on a 9-point Likert scale (1: strongly disagree, through 9: strongly agree). We chose 80% agreement as the cut-off for consensus based on findings from a systematic review of Delphi studies^13^. We considered consensus to be reached if 80% of respondents scored the question within the top third (score 7 to 9 to include) or bottom third (score 1 to 3 to exclude) of the 9-point scale.

**Round 1**. Participants ranked the importance of all questions via the online survey. We asked participants for any additional comments they wished to provide on each question using free text boxes.

**Round 2**. Based on the results and comments from round 1, the research team removed questions that reached consensus, eliminated or modified ambiguous questions and included additional questions driven by comments from participants. For example, we received suggestions from several participants proposing that we adjust the question on collaborator roles and their ranked importance in helping to solve the problem of predatory journals. As a result, we added two additional collaborator roles that could be ranked: researchers and academic societies. We then invited participants to complete round 2 of the Delphi. In the round 2 survey invitation, we provided participants with summarized, de-identified results from round 1: a narrative summary of the survey results along with measures of central tendency (weighted average) and dispersion (range) summarized for each question. One participant requested the original comments from round 1, which we then provided. We asked participants to again rate the importance of the remaining survey questions, using the same scale as in round 1 and referring to the results provided from round 1. Text boxes were again used to solicit additional comments.

**Round 3**. Participants were invited to attend our Predatory Summit to complete round 3 of the Delphi. Results from the first two rounds were available to attendees prior to the event (April 19-20, 2019 in Ottawa, Canada) on the Open Science Framework (https://osf.io/46hwb/). We encouraged attendees to look over the summarized results, which included measures of central tendency (weighted average), dispersion (range), and comments provided by participants for each question. A final round of voting was held in person at the Summit for questions that had not reached consensus using Poll Everywhere (https://www.polleverywhere.com/). Participants could observe results in real-time as data were collected. For this round, we used a 3-point Likert scale that included the same 9 original responses in a simplified format (1 = 1-3 = strongly disagree; 2 = 4-6 = neutral; 3 = 7-9 = strongly agree). Face-to-face presentations and discussions took place at the Summit to further refine, contextualize and finalize the results (see Summit agenda: https://osf.io/thsgw/).

### Participants

Authors (group 1): A previous scoping review identified 344 articles that discussed predatory journals^9^. From these articles, we identified the corresponding authors, removed any duplicates, extracted author contact information, removed any authors whose contact information was not available, and sent an invitation to the remaining 198 authors to complete round 1 of our survey. Summit invitees and participants (group 2): Through snowball and purposive sampling of targeted experts, we identified 45 noted experts in predatory journals and journalology to participate in the Delphi process and to attend our Predatory Summit. Invitees and participants were international experts representing the varied stakeholders affected by predatory journals, including funders, academic institutions, librarians and information scientists, digital scientists, researchers involved in studying predatory journals, legitimate journals, and patient-partners. Two individuals had planned to attend the Summit and so participated in rounds 1 and 2 of the Delphi, but did not attend the Summit and so could not participate in round 3.

### Statistical Analysis

We reported discrete variables as counts/proportions. Continuous variables were reported as medians and ranges.

## RESULTS

### Deviations from our protocol

We did not deviate from the study procedures outlined in our protocol.

### Comparing round 1 results between groups 1 and 2

The round 1 Delphi results of groups 1 (authors) and 2 (Summit invitees and attendees) were similar, with agreement on consensus or no consensus on 30 out of 35 questions. The five remaining questions reached consensus on inclusion for the Summit invitees and participants (group 2) but not for authors (group 1) (Appendix B). Descriptions of the discrepancies between groups on these five items are also briefly detailed in the results below (see detailed results from round 1, group 1 here: https://osf.io/vmura/, round 1 group 2 here: https://osf.io/sry9w/; see https://osf.io/d5463/ for a complete comparison between results of both groups, highlighting which questions had responses that differed by more than 10% between groups).

For reasons of feasibility and because of the similar results between groups, as indicated in the study protocol, we invited only the Summit invitees and participants (group 2) to continue with rounds 2 and 3. We report results of only the Summit invitees and participants (group 2) as respondents going forward.

### Respondent Demographics

Twenty-one of 45 Summit invitees and participants identified as female (47%, Table 1). There was international representation including participants from lower-middle income economies (India: n = 1, 2%), and upper-middle income economies (South Africa: n = 4, 9%). Summit invitees and participants reported representing a variety of stakeholder groups, with some individuals representing more than one group, including researchers (n = 22, 49%), funders (n = 13, 29%), policy makers (n = 2, 4%), journal editors (n = 5, 11%) and patient partners (n = 2, 4%).

**Table 1.** Respondent characteristics

| **Characteristics** | **N (%)** |
| --- | --- |
| Gender |  |
| Female | 21 (47) |
| Male | 24 (53) |
| Stakeholder group* |  |
| Academic institution | 4 (9) |
| Funder | 13 (29) |
| Government | 1 (2) |
| Journal Editor | 5 (11) |
| Patient partner | 2 (4) |
| Policy maker | 2 (4) |
| Publisher | 3 (7) |
| Research network | 2 (4) |
| Researcher | 22 (49) |
| Student | 1 (2) |
| Other | 1 (2) |
| Geographic location |  |
| Canada | 24 (53) |
| India | 1 (2) |
| Italy | 3 (7) |
| Netherlands | 1 (2) |
| South Africa | 4 (9) |
| Sweden | 1 (2) |
| Switzerland | 4 (9) |
| UK | 3 (7) |
| USA | 2 (4) |
| International | 2 (4) |

\*Percentages do not add up to 100 since some participants identified as part of more than one stakeholder group.

### Participation by round

Of the 45 survey invitation emails sent for round 1 of the Delphi, 35 invitees completed the survey (83%). In round 2, 32 completed the survey (76%). In both rounds, participants included detailed comments in the free text boxes, supporting their responses or describing additional considerations on the topic, for each of the questions. Of the 43 participants who met face-to-face at the Predatory Summit, we received responses from 30 to 38 participants for each question (70-88%). The variance in response rates at the Summit could have been due to participants stepping out of the room during a question, arriving late, or preferring not to comment on all items. A summary of all items reaching consensus, and the round at which consensus was reached, can be found in Table 2.

**Table 2.** Delphi items to reach consensus as very important or strongly supported

| Delphi Items | Round when consensus reached | N (%) |
| --- | --- | --- |
| 1. How important is it to develop a consensus definition for predatory journals? | 1 | 31 (94) |
| 2. **Characteristics** that differentiate predatory and legitimate journals: |  |  |
| 2a. The journal's operations are deceptive (i.e. misleading; not truthful) | 1 | 33 (94) |
| 2b. The journal's operations are not in keeping with best publication practices (e.g. no membership in COPE) | 1 | 28 (80) |
| 2c. Journal has low transparency regarding its operations | 1 | 28 (80) |
| 2d. Fake impact factors are promoted by the journal | 1 | 33 (94) |
| 3. **Markers** that best differentiate predatory and legitimate journals: |  |  |
| 3a. The journal has no retraction policy | 3 | 36 (95) |
| 3b. The journal solicits manuscripts through aggressive or persuasive emails | 1 | 32 (91) |
| 3c. The contact details of the publisher are not easily verifiable | 1 | 34 (97) |
| 4. **Empirically derived data** that best differentiate predatory and legitimate journals: |  |  |
| 4a. The journal does not mention a Creative Commons license | 1 | 28 (80) |
| 4b. The journal's homepage has a 'look and feel' of being unprofessional | 1 | 30 (86) |
| 4c. Editors and editorial board affiliations with the journal are not verifiable | 1 | 35 (100) |
| 4d. The journal is not a member of COPE | 1 | 28 (80) |
| 5. Should public funders fund research about predatory publishing? | 1 | 28 (80) |
| 6. Several groups have developed checklists to help authors identify and avoid predatory publishers. Should a **single, coherent checklist** be developed to replace existing checklists? | 2 | 25 (83) |
| 7. Is there merit in developing resources or **educational materials** regarding predatory journals / publishers **in languages other than English?** | 3 | 26 (87) |
| 8. Should efforts be made to differentiate predatory journals from very low quality journals? | 1 | 30 (86) |
| 9. Is there merit in developing a '**one stop shop**' website to consolidate information, training and educational materials about predatory journals? | 1 | 28 (80) |
| 10. Is there merit in establishing a **predatory journal research observatory?** | 2 | 24 (80) |

Below we review the Delphi results for each question, within each of the three categories of questions (see Appendix A for complete results):

### 1. Definition of predatory journals

#### Importance of developing a consensus definition for predatory journals

Consensus was reached in round 1 on the need to develop a consensus definition of predatory journals (n = 32, 94%).

#### Should the term “predatory” be changed?

There was no consensus on whether the term ‘predatory’ should be changed. Respondents were almost equally split across all lateral thirds of the Likert scale (no name change: n = 10, 29%; neutral: n = 13, 37%; alternative name required: n = 12, 34%). Round 2 results were similarly divided across the scale. In round 3, after in-person discussion, consensus was not reached during live voting.

#### What alternative name(s) would you suggest?

Consensus on an alternative name was not reached in either of the first two rounds from among the following terms: dark journals / publishers; deceptive journals / publishers; illegitimate journals / publishers; or journals / publishers operating in bad faith. In rounds 1 and 2, many respondents agreed that *dark journals / publishers* was a “terrible name” (n = 21, 63%; n = 20, 67%). The name with the greatest positive traction in both rounds was *deceptive journal / publisher* (n = 25, 71%; n = 20, 67% thought this was an “excellent name”).

After not reaching consensus in round 3 on the question of a name change, participants discussed the merits and challenges of this task. Some reasons in support of a name change included the association of predatory with the idea that the author is always a victim of a predatory journal/publisher. However, some authors publish in predatory journals knowing that the journal is predatory, for ease of publication^14^. Other reasons to not use the term predatory, as was discussed at the Summit, include its affiliation with the Beall’s list and the fact that other terms may be more descriptive, such as the term “deceptive”.

Participants discussed the challenges associated with changing an established term, including challenges in identifying literature, disseminating and promoting the new name internationally, and updating existing educational materials and funder statements.

At the Summit, it was concluded that changing an already established term would likely be confusing to the scientific community and not in the best interest of moving this agenda forward. It was recommended that the term “predatory” continue to be used and that limitations to the term, as indicated above, be recognized^10^.

#### Characteristics that differentiate between predatory and legitimate journals

Respondents were asked to rate the importance of four different characteristics in identifying the journal as predatory. We defined characteristics as distinct features of all predatory journals.

These characteristics are unique to predatory journals and generally do not occur at legitimate high-quality open access journals. Consensus was reached for all four of the following: the journal’s operations are deceptive; the journal’s operations are not in keeping with best publication practices (e.g. no membership in COPE) (for this item, results from group 1 (authors) were similar to group 2 (Summit invitees and participants), however, group 2 did not reach consensus (67% thought this was *a very important characteristic*)); the journal has low transparency regarding its operations; fake impact factors are promoted by the journal.

#### Markers or distinguishing features that differentiate between predatory and legitimate journals

Respondents were asked to rate the importance of seven different markers in identifying a journal as predatory. We defined markers as features that are *common* among predatory journals. Not all markers are present in all predatory journals. Markers may be considered “red flags” of poor journal quality. There was consensus in round 1 that two of the seven markers were very important in identifying predatory journals: the journal solicits manuscripts through aggressive or persuasive emails; and, contact details of the publisher are not easily verifiable. The remaining five questions did not reach consensus in round two: the journal promises a very quick peer review and turn around; the journal promises rapid publication; the journal has no retraction policy – this question was missed in round 2, in error – in round 1 it almost reached consensus with 79% of respondents rating this as a very important marker; the journal is not a member of COPE; the journal is not listed in the DOAJ. In round 3, not having a retraction policy reached consensus as a very important marker in distinguishing between a predatory journal and a legitimate one.

#### Empirically derived data that differentiate between predatory and legitimate journals

Respondents were asked to rate the importance of six types of empirically derived data in identifying the journal as predatory. We defined ‘empirically derived’ data as data resulting from experiments or statistical analyses that indicate differences between predatory journals and legitimate open access journals/publishers^2^. In round 1, consensus was reached on four of the six questions, indicating very important data elements in identifying a predatory journal: the journal’s homepage has a ‘look and feel’ of being unprofessional; editors and editorial board affiliations with the journal are not verifiable; the journal is not a member of COPE; the journal does not mention a Creative Commons (CC) license. For this last item (journal does not mention a CC license), results from group 1 (authors: 43% thought this was a *very important characteristic*) differed from group 2 (Summit invitees and participants: 80% (consensus reached) thought this was *a very important characteristic*). This discrepancy could be due to the fact that Summit participants, including three journal publishers and five journal editors, would be more knowledgeable about the nuances of a CC license). The remaining two questions did not reach consensus in rounds 2 or 3: the journal’s article processing charge (APC) is considerably lower than legitimate OA journals; the journal is not listed in the DOAJ.

### 2. Educational outreach and policy initiatives on predatory publishing

#### Should public funders fund research about predatory publishing?

In round 1, consensus was reached that public funding is essential to study and address the issue of predatory publishing (n = 28, 80%). Although the group of authors (group 1) did not reach consensus on this item, their responses suggest a response similar to the Summit invitees and participants (72% of authors thought that *funding is essential*).

#### Should research published in predatory journals be included in systematic reviews and meta-analyses

In round 1, consensus was not reached on whether research published in predatory journals should be included in systematic reviews or meta-analyses. The research group decided to remove this question from the survey after considering the fact that respondents are not experts in systematic review or meta-analysis methodology, and therefore would not be well-positioned to evaluate this item.

#### Do multiple checklists available for assessing predatory journals confuse prospective authors?

Consensus was not reached in any of the three rounds to determine if this was or was not a *serious problem*.

#### Should a single, coherent checklist should be developed to replace existing checklists?

There was consensus in round 2 that a single checklist should be developed (n = 25, 83%).

#### Importance of referencing and promoting pay-to-access lists indicating good quality journals and other lists indicating potential predatory journals

Questions on the good quality lists and lists of potential predatory journals did not reach consensus in any of the three Delphi rounds. In rounds 1 and 2, half of the participants (n = 17, 50%; n = 17, 50%) thought it was *very important* to reference and promote both types of lists. In round 3, there was a switch, and more participants thought that referencing and promoting lists of potential predatory journals was more important (n = 21, 58%) than referencing and promoting pay-to-access lists of good quality journals (n = 7, 23%). The change in voting could have been due to discussions at the Summit regarding pay-to-access lists as counter to the principles of open access and equity. These discussions could have been influenced as well by the presentation by Michaela Strinzel and Anna Severin (both from the Swiss National Science Foundation), delivered at the Summit, demonstrating the overlap between lists of good quality journals and lists of potential predatory journals^15^.

#### Ranking the level of importance of collaborators in helping solve the problem of predatory journals

In round one, six collaborators were named and participants ranked them in order of importance: 1- Academic institutions; 2- Funders; 3- Libraries; 4- COPE; 5- Journals / publishers; 6- DOAJ. In this round, participants commented on other potential collaborators, many of whom suggested researchers and academic societies. These two categories of collaborators were added in round 2. The ranking changed slightly in this round, with the new additions, as follows: 1- Academic institutions; 2- Researchers; 3- Journals / publishers; 4- Funders; 5- Libraries; 6- Academic societies, e.g. learned societies; 7- COPE; 8- DOAJ;. Since this question did not require consensus, it was not repeated in round 3.

#### Merit in developing resources or educational materials regarding predatory journal / publishers in languages other than English

This question almost reached consensus as an *excellent idea* in the first two rounds (n = 27, 77%; n = 23, 77%). The question then reached consensus in round 3 (n = 26, 87%). Participants across the first two rounds suggested translation to other languages including French, Spanish, Indian languages (Hindi, Bengali), German, Chinese (Mandarin) and Arabic, among others.

#### Strategies that would be best suited to solve the challenge of predatory journals faced by researchers in low and middle income countries (LMIC)^1^

Participants were asked to check options that they felt were suitable strategies. Two strategies received high response rates in round 1: A checklist to help detect predatory journals (n = 26, 72%); and a “One stop shop” website that consolidates information, training, and education about predatory journals / publishers (n = 30, 83%). An error in one of the strategies listed may have contributed to false results in both rounds 1 and 2. That strategy option should have read: “Paywalled whitelists that name trustworthy or legitimate journals” however, it read: “Paywalled whitelists that name predatory journals / publishers”. There could have been confusion about this strategy option since whitelists in this context typically include legitimate or trustworthy journals, and not potential predatory journals or ones to avoid. In rounds 1 and 2, the journal authenticator^2^ received high response rates as well (n = 21, 58%; n = 23, 77%). Comments from participants in the two rounds included other suggested strategies, for example, moving away from a “publish or perish” culture in academia which addresses the demand side of predatory journals rather than the supply side; more support for ambassadors (e.g. at the DOAJ) and the International Network for the Availability of Scientific Publications (INASP) workshops onsite; and a number of others indicated that they are not experts in the needs of communities in LMICs. Consensus was not relevant for this question and it therefore was not included in round 3.

#### Should efforts be made to differentiate predatory journals from very low quality journals?

There was consensus in round 1 that important efforts should be made to differentiate between predatory journals and journals of very low quality (n = 30, 86%). Although the group of authors (group 1) did not reach consensus on this item, their responses suggest a response similar to the Summit invitees and participants (77% of authors thought that *important efforts should be made*). By very low quality we mean journals that are under-resourced, or are run by an editorial board that is uninformed. These journals would not be considered predatory, however, their practices are still well below accepted publication science standards.

### 3. Developing technological solutions to stop submissions to predatory journals and other low-quality journals

#### Is there merit in developing a ‘one stop shop’ website to consolidate information, training and educational materials about predatory journals?

Consensus was reached in round 1 that a ‘one stop shop’ was an excellent idea (n = 28, 80%). Although the group of authors (group 1) did not reach consensus on this item, their responses suggest a response similar to the Summit invitees and participants (76% of authors thought that developing a ‘one stop shop’ is an *excellent idea*).

#### Is there merit in developing a journal authenticator^2^?

There was support in all three rounds for the development of a journal authenticator (n = 26, 74%; n = 23, 77%; n = 27, 79%), however, this question did not reach consensus.

#### Is there merit in establishing a predatory journal research observatory^3^?

Consensus was reached in round 2 that there is strong support in establishing a predatory journal research observatory (n = 24, 80%).

## DISCUSSION

We conducted a modified Delphi with the aim of generating a consensus definition of predatory journals, as well as consensus on how the research community should respond to predatory journals. We came to consensus on 18 survey items out of a total of 33 (not including the question on inclusion of data in systematic reviews removed after round 1) (see Table 2). These consensus items included the characteristics, markers and empirically derived data to be included in the definition of predatory journals and publishers.

In-person deliberations at the Summit proved to be an important step in coming to consensus on the decision not to change the term ‘predatory’. Lengthy discussions among Summit participants centred on establishing a term that best described the activities of predatory journals and publishers, while weighing the challenges of a change in an established term. The group concluded that any change in terminology would hinder the efforts of the scholarly community to stop publication in predatory journals, and recommended continuing to use the term ‘predatory’.

We were able to reach consensus on avenues of educational outreach and policy initiatives, agreeing that public funds should be allocated to research about predatory publishing, and that a single checklist should be developed to help authors detect predatory journals (see systematic review of checklists to detect predatory journals^16^). Resources such as these should be developed in languages other than English. Some agreed-upon strategies to address the problem of predatory journals and publishers in low- and lower-middle income economies include: a checklist to detect predatory journals, a ‘one-stop-shop’ website, and a journal authenticator. We agreed that various collaborators have important roles in moving this agenda forward, including those identified as most responsible: academic institutions, researchers and journals and publishers. Finally, we reached consensus that important efforts were necessary to distinguish very low quality journals from predatory journals.

Future directions suggested included the development of technological solutions to stop submissions to predatory journals and other low-quality journals. We reached consensus on developing a ‘one-stop-shop’ website to consolidate information, training and educational materials about predatory journals and establishing a predatory journal research observatory.

The Delphi results have since been used to inform the development of a consensus statement on predatory journals and to map next steps in addressing predatory journals^10^. With this consensus definition and a roadmap for future action, we are now better positioned to study the phenomenon of predatory journals / publishers, more precisely inform policy and education initiatives, and direct resources appropriately.

### Limitations

The findings of this modified Delphi study are limited by the fact that only selected participants contributed to the survey results. Inclusion of a larger number of individuals with different expertise and backgrounds may have changed the results. We attempted to be comprehensive in the development of the survey questions; however, in compiling the final list, some questions may have been overlooked. A final limitation that may have changed the survey outcomes are possible issues with language not being preserved within the original scoping review from which we developed survey questions, or nuances in language not being captured in questions.

## CONCLUSION

Bringing together international participants representing diverse stakeholder groups allowed for a comprehensive synthesis of survey responses to inform the development of a definition of predatory journals and publishers. The Delphi identified characteristics of predatory journals and publishers, education outreach and policy initiatives as well as guidance on future directions and the development of technological solutions to stop submissions to predatory journals and other low-quality journals.

## Data Availability

All data are available upon request.

https://osf.io/z6v7f/

https://osf.io/ysw3g/

https://osf.io/46hwb/

https://osf.io/thsgw/

https://osf.io/vmura/

https://osf.io/sry9w/

https://osf.io/d5463/

## Acknowledgements

We would like to thank the following individuals at the Ottawa Hospital Research Institute for their help in reviewing and piloting the Delphi survey: Andrew Beck, Matthew McInnes, Danielle Rice and Beverly Shea.

We would like to the following individuals who participating in the Delphi survey: Kristiann Allen, Katherine Akers, Clare Ardern, Tiago Barros, Monica Berger, Pravin Bolshete, Alison Bourgon, Marion Broome, MP Cariappa, Imti Choonara, Mary M. Christopher, Marco Cosentino, Tammy Clifford, Lucia Cugusi, Lenche Danevska, Sergio Della Salla, Mahash Devnani, Helen J. Dobson, Michael Donaldson, Stefan Eriksson, Matthias Egger, Kevin Fitzgibbons, Danielle Gerberi, Vinicius Giglo, Seethapathy Gopalakrishnan Saroja, Ian D. Graham, Martin Grancay, Farokh Habibzadeh, Evelym Hermes-Desantis, Fang Hua, Matt Hodgkinson, Shobhit Jain, Hamid R. Jamali, Manthan Janodia, Scott Kahan, Nolusindiso (Sindi) Kayi, Shawn Kennedy, SC Lakhotia, Mahlubi (Chief) Mabizela, Andrea Manca, Alexandre Martin, Ana Marusic, Stuart McKelvie, Aamir Memon, Eric Mercier, Katrin Milzow, Johann Mouton, Marilyn Oermann, Tom Olyhoek, Alexander Ommaya, Ayokunle Omobowale, Cetin Onder, Sanjay Pai, Kishor Patwardhan, Alexandru Petrisor, Durga Prasanna Misra, Filipe Prazeres, Laurie Proulx, Regina Reynolds, Jason Roberts, Marc Rodger, Kem Rogers, Anna Severin, Michaela Strinzel, Mauro Sylos-Labini, Marthie van Niekerk, Jelte M. Wicherts, Roger Wilson, Tim Wilson, Riah Wiratningsih, Susan Zimmerman.

## Appendix A. Delphi results by round, N (%)

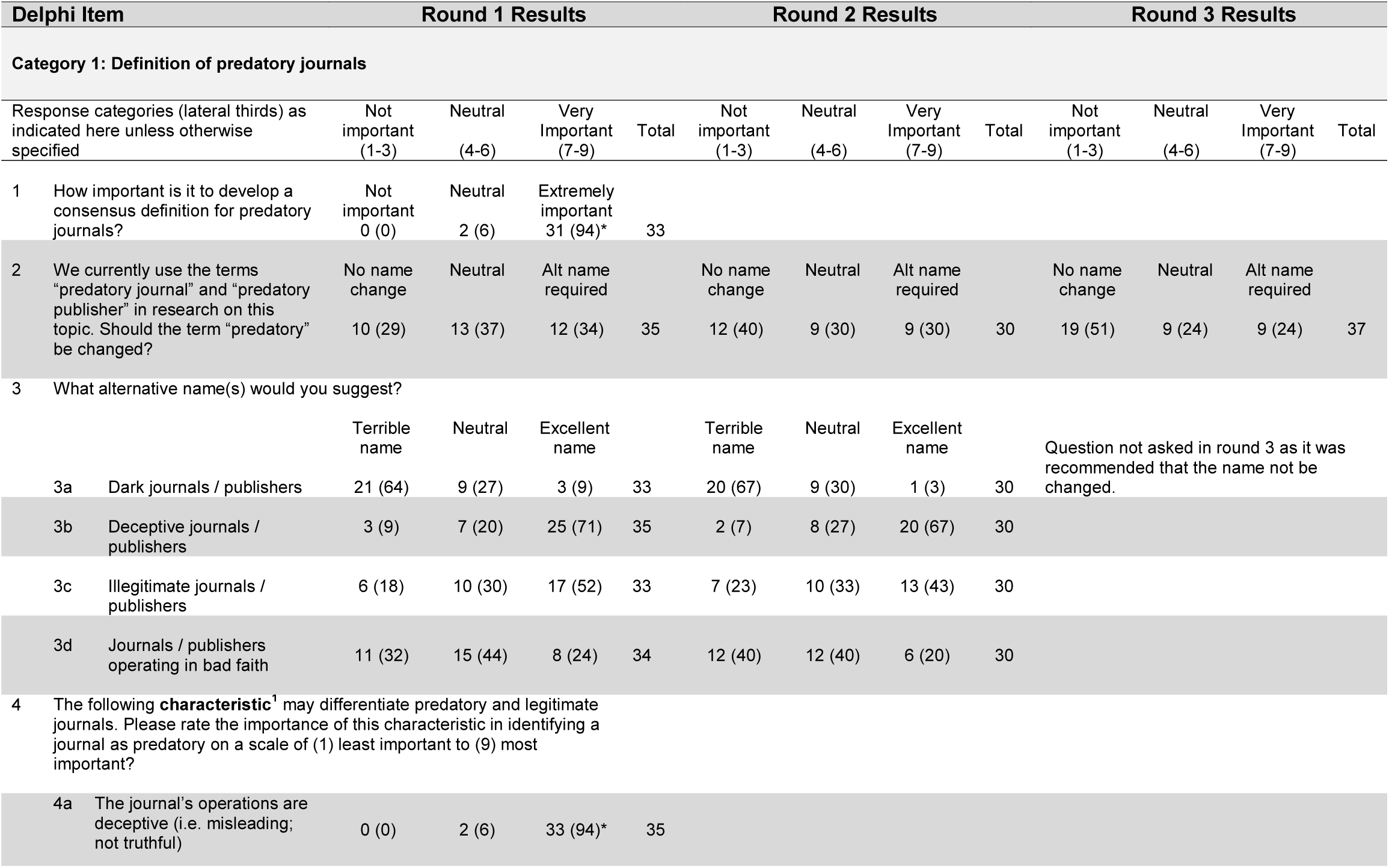
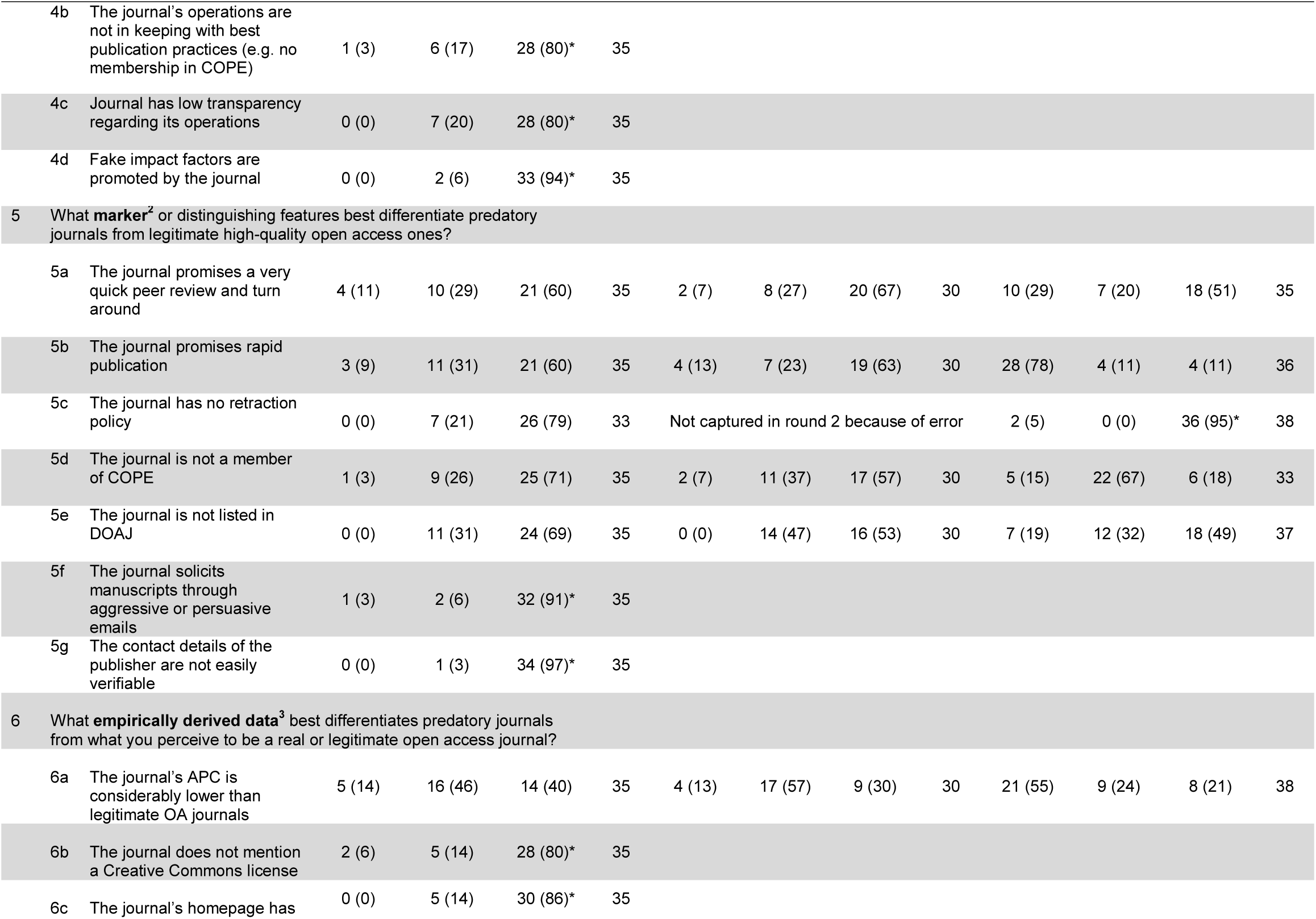
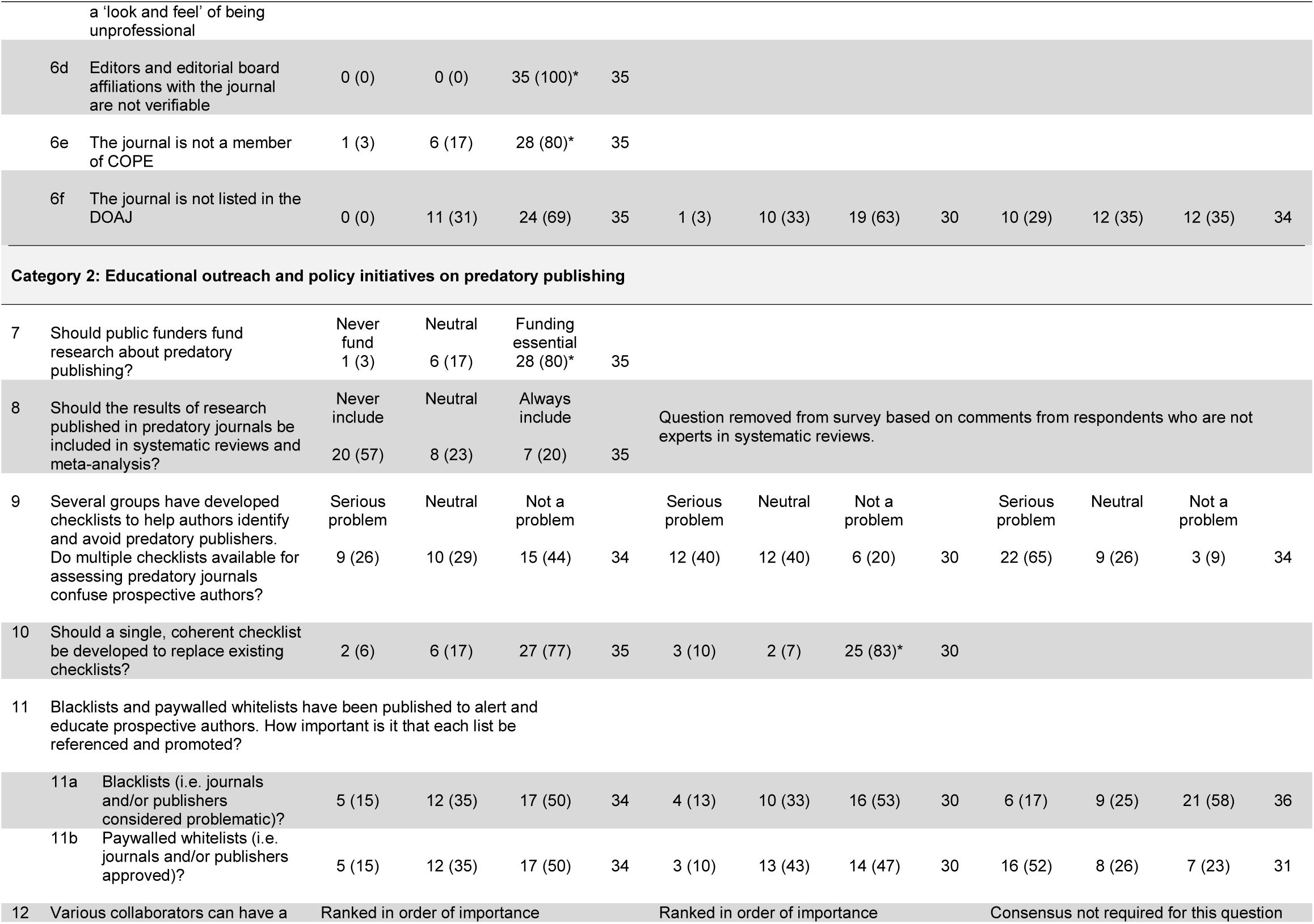
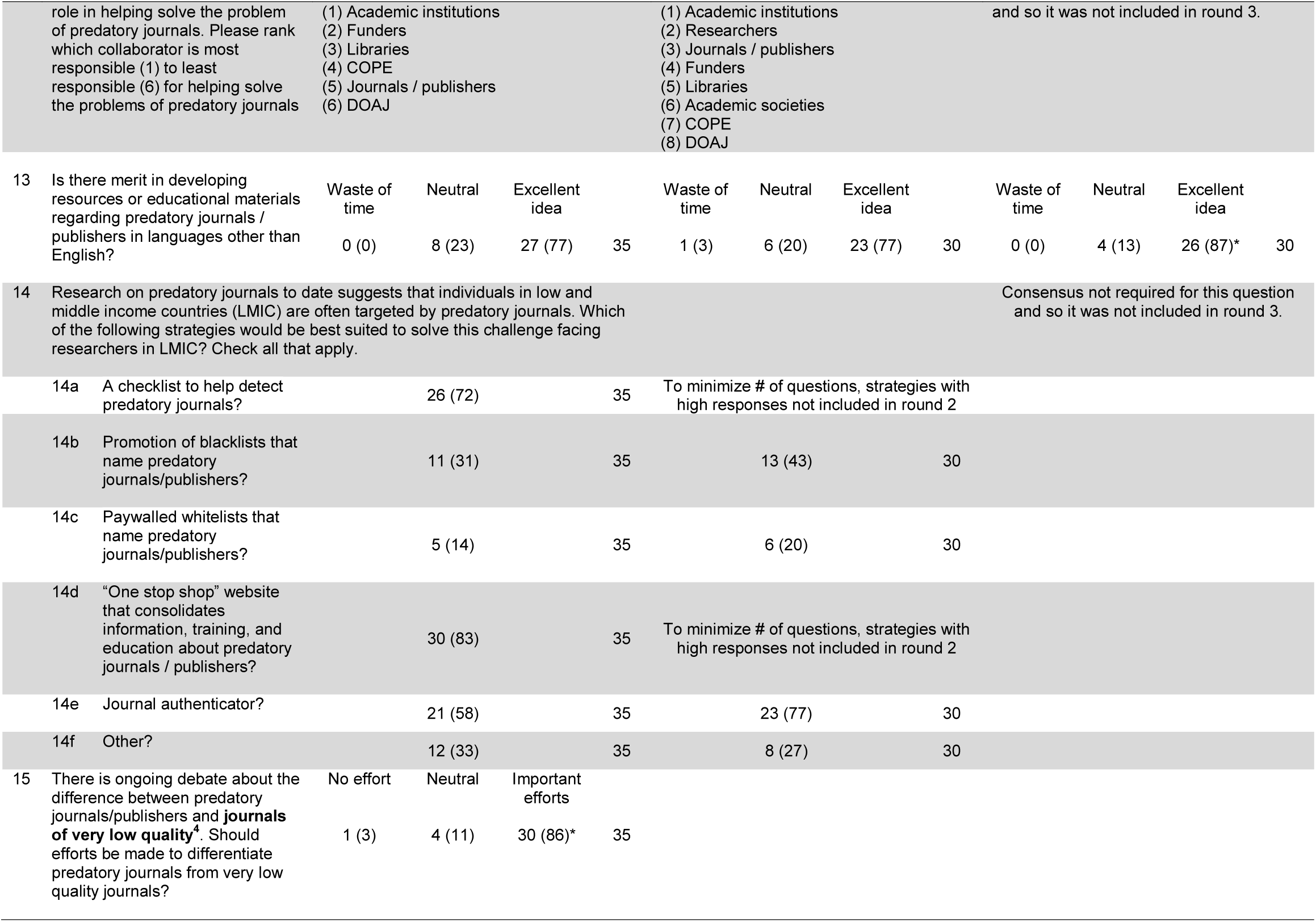
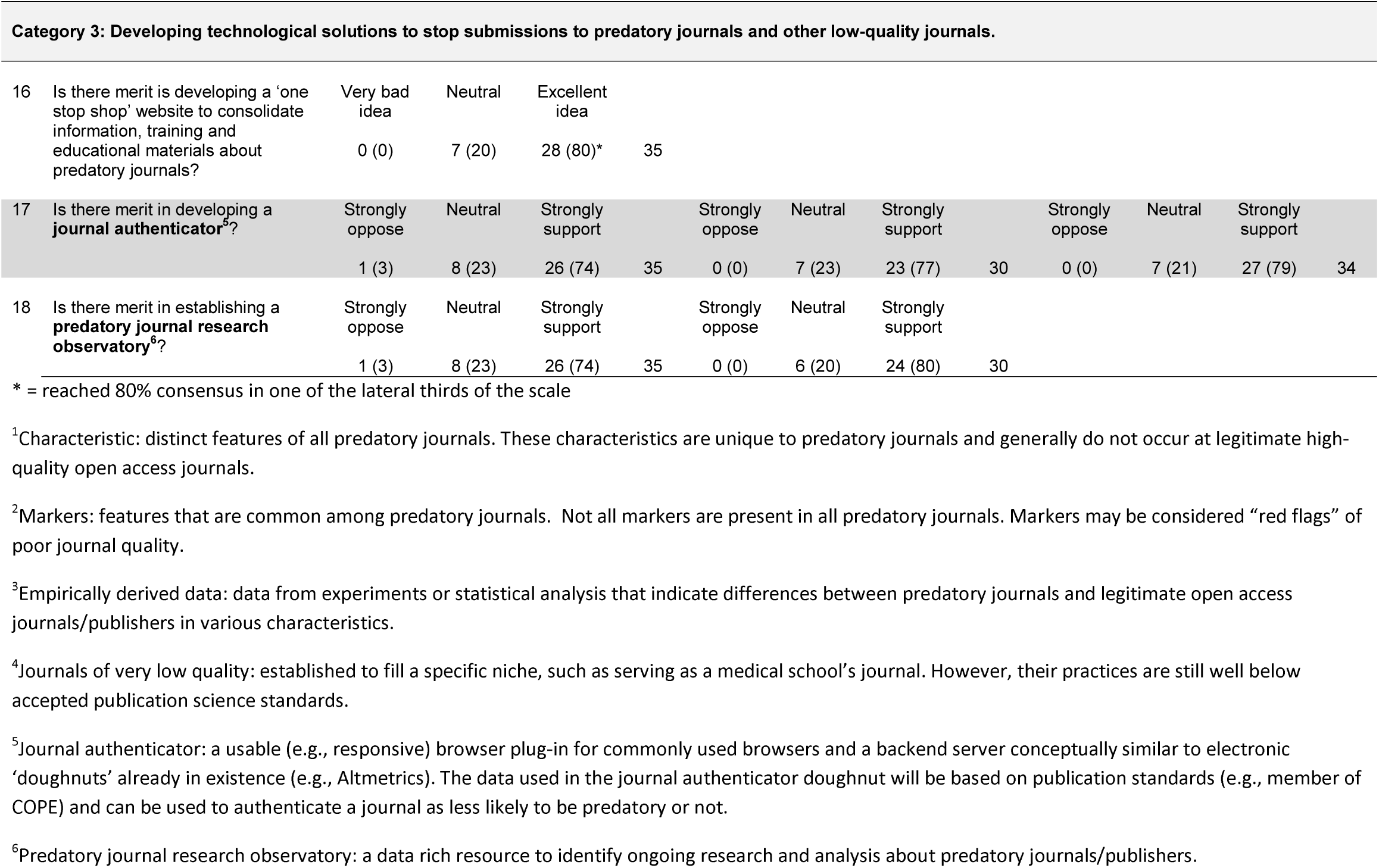

## Appendix B. Differences in consensus results for round one between authors identified in scoping review by Cobey et al., (2018) (n = 72) (group 1) and Summit invitees and participants (n = 45) (group 2)

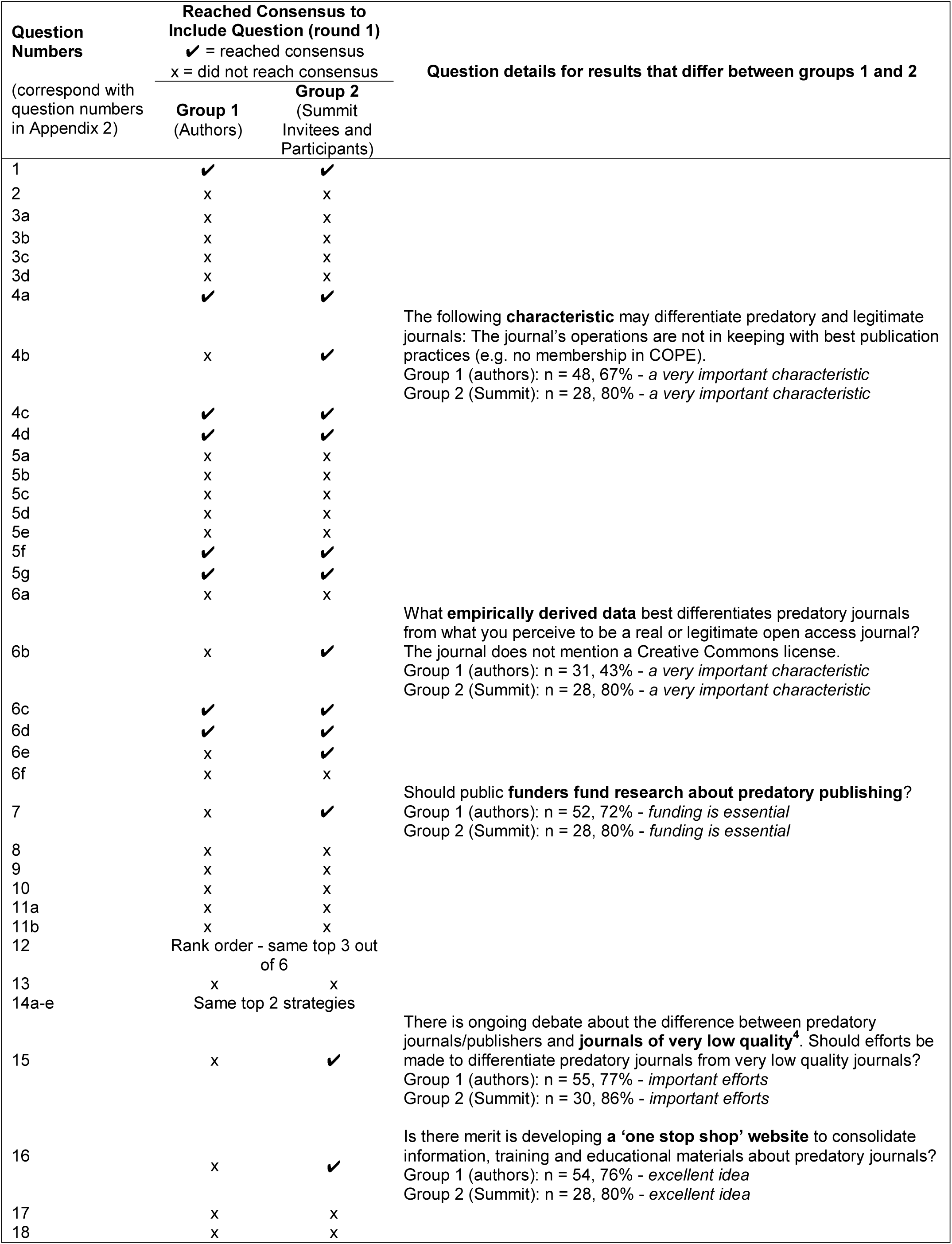

## Footnotes

1 Currently, the World Bank uses new classifications: low-income, lower-middle-income, upper-middle-income and high-income economies.

2 A usable (e.g., responsive) browser plug-in for commonly used browsers and a backend server conceptually similar to electronic ‘doughnuts’ already in existence (e.g., Altmetrics). The data used in the journal authenticator doughnut will be based on publication standards (e.g., member of COPE) and can be used to authenticate a journal’s quality status.

3 A data rich resource to identify ongoing research and analysis about predatory journals/publishers

